# Associations between transdiagnostic psychological processes and global symptom severity among outpatients with various mental disorders: A cross-sectional study

**DOI:** 10.1101/2024.06.24.24309393

**Authors:** Anna Katharina Frei, Thomas Studnitz, Britta Seiffer, Jana Welkerling, Johanna-Marie Zeibig, Eva Herzog, Mia Maria Günak, Thomas Ehring, Keisuke Takano, Tristan Nakagawa, Leonie Sundmacher, Sebastian Himmler, Stefan Peters, Anna Lena Flagmeier, Lena Zwanzleitner, Ander Ramos-Murguialday, Sebastian Wolf

## Abstract

**Objective:** Knowledge about transdiagnostic factors associated with global symptom severity among patients diagnosed with various mental disorders remains limited. This study examined the cross-sectional associations between transdiagnostic psychological processes (emotional regulation, emotional intelligence, sleep quality, perceived stress, fear of the coronavirus, repetitive negative thinking), amount of physical activity, and sedentary behavior with global symptom severity, while controlling for sociodemographic data (age, gender, employment status, relationship status, educational level).

**Methods:** Data from 401 outpatients, aged 42.08 years on average (SD = 13.26; 71.3% female), diagnosed with depressive disorders, non-organic primary insomnia, agoraphobia, panic disorder, and/or posttraumatic stress disorder were analyzed. Data was collected from 10 different study sites between March 2021 and May 2022 for cross-sectional analysis. The influence of predictors of global symptom severity was determined using multiple regression. Global symptom severity was measured using the Global Severity Index, derived from the Brief Symptom Inventory-18. Predictors were measured using validated scales and physical activity was additionally assessed via accelerometer-based sensors.

**Results:** The proposed model explained 40.3% of variance on the Global Severity Index. Higher levels of repetitive negative thinking, fear of coronavirus, perceived stress, worse sleep quality as well as greater difficulties with emotional regulation and emotional intelligence were significantly associated with higher global symptom severity in our clinical outpatient sample; however, no associations were found for physical activity, and sedentary behavior.

**Conclusion:** Transdiagnostic psychological processes were significantly associated with global symptom severity. Additional prospective longitudinal studies with transdiagnostic samples are necessary to explore possible causal relationships.

**Key Practitioner Message:**

- Repetitive negative thinking, perceived stress, sleep quality, and emotional regulation can be considered distinct and individual transdiagnostic psychological processes associated with global symptom severity among outpatients diagnosed with various mental disorders.
- Treatment approaches aiming at improving transdiagnostic predictors of various mental disorders might be an efficacious target in reducing global symptom severity.
- Among outpatients (75% already receiving psychological or pharmacological treatment) diagnosed with depressive disorders, non-organic primary insomnia, agoraphobia, panic disorder, and/or PTSD, sociodemographic factors appear not to be predictive for global symptom severity.

## 1. Introduction

Major depressive disorder, anxiety disorders (such as panic disorder and agoraphobia), posttraumatic stress disorder (PTSD) and non-organic primary insomnia exhibit high comorbidity (Kessler, Chiu, Demler, Merikangas, & Walters, 2005; McGrath et al., 2020) and are characterized by partial symptom overlap (e.g., sleep problems) (American Psychiatric Association, 2022). Several factors have been found that influence these mental disorders by posing a risk for their onset, predicting their symptom severity and/or contributing to their maintenance. Traditionally, variables involved in the development and maintenance of psychopathology have typically been studied from a disorder-specific perspective. However, there is increasing evidence that transdiagnostic processes play an important role across different diagnostic categories (Dalgleish, Black, Johnston, & Bevan, 2020).

Depression, anxiety disorders, PTSD, and insomnia are often referred to as "stress-related" disorders (Palagini et al., 2023; Smoller, 2016), as severe acute stressful life events or prolonged exposure to stressors are risk factors for the development and exacerbation of psychiatric symptoms (Healey et al., 1981; Kessler, 1997; Moreno-Peral et al., 2014; Schwarzer & Luszczynska, 2012). The COVID-19 pandemic characterized by high mortality rates, isolation and overwhelmed healthcare systems can be considered such stressful life event, or rather period, and elevated stress levels during that time were reported (Gamonal-Limcaoco et al., 2022). Indeed, meta-analytic results suggest that fear of the coronavirus is related to anxiety, traumatic stress, depression and insomnia (Simsir, Koc, Seki, & Griffiths, 2022).

Emotional regulation can be considered a coping style for stress by managing typical stress emotions, such as anger, anxiety, or shame. Contrary, deficits in emotional regulation favor the development of depression (Berking, Wirtz, Svaldi, & Hofmann, 2014) and among patients suffering from depression these deficits are seen as one key factor in the maintenance of symptoms (Ehring, Tuschen-Caffier, Schnulle, Fischer, & Gross, 2010). Impaired emotional regulation also seems to be significantly correlated with higher symptom severity of PTSD (Ehring & Quack, 2010). Emotional intelligence, a specific personality trait that involves an individual’s self-perceived ability to understand, process and regulate emotional information in oneself and others (Petrides, Furnham, & Mavroveli, 2007; Petrides, Pita, & Kokkinaki, 2007) is associated with severity of insomnia symptoms (Emert, Tutek, & Lichstein, 2017).

Repetitive negative thinking is often conceptualized as a maladaptive coping strategy for distress as it tends to impede effective problem-solving abilities (Ward, Lyubomirsky, Sousa, & Nolen-Hoeksema, 2003). It has been found to be present in a range of mental disorders, including depression, PTSD, insomnia, and anxiety disorders (Ehring & Watkins, 2009) and to be a risk factor for higher severity and maintenance of depressive and anxiety-related symptoms (Spinhoven, van Hemert, & Penninx, 2018). In addition, a systematic review about rumination as a subtype of repetitive negative thinking concludes that it predicts PTSD symptom severity among patients diagnosed with PTSD (Moulds, Bisby, Wild, & Bryant, 2020).

Some individuals particularly engage in repetitive negative thinking at bedtime, which has been linked to reduced subjective sleep quality (Takano, Iijima, & Tanno, 2012) and shortened sleep duration (Nota & Coles, 2018). Nonetheless, adequate sleep is essential for effectively managing stress (Hamilton, Catley, & Karlson, 2007). Therefore, establishing and maintaining proper sleep routines or implementing sleep hygiene practices may serve as coping mechanisms for stress. Importantly, sleep disturbances are not only part of the symptom criteria for many different disorders (e.g., depression, PTSD) but are also associated with higher levels of psychiatric symptom severity among patients with mood and anxiety-related disorders, and PTSD (Belleville, Guay, & Marchand, 2009; Kallestad et al., 2012).

Finally, physical activity can buffer the negative effects of stress in daily life (Hachenberger et al., 2023) and has been identified as an important lifestyle variable related to psychological well-being among people with mental disorders (Firth et al., 2020). However, meta-analytic findings suggest that suffering from mental disorders is associated with reduced involvement in physical activity and elevated levels of sedentary behavior (Schuch et al., 2017; van den Berk-Clark et al., 2018; Vancampfort et al., 2017). Importantly, sedentary behavior is associated with an increased risk of depression (Huang et al., 2020), anxiety (i.e., any anxiety disorder or anxiety symptoms) (Allen, Walter, & Swann, 2019) and insomnia (Yang, Shin, Li, & An, 2017). In fact, individuals who use physical activity as a strategy for affect regulation showed fewer psychiatric symptoms when being physically active compared to inactive individuals (Rösel et al., 2022). Furthermore, in people with mental disorders, those who show higher levels of daily physical activity are more likely to have better global functioning (Derhon, Guimaraes, Vancampfort, Moraleida, & Schuch, 2023). However, there is a lack of studies examining the impact of daily physical activity on mental health and meta-analytic findings suggest that leisure-time physical activity is related to positive mental health outcomes, but the associations with other types of activity, such as transportation or household chores, remain inconsistent (White et al., 2017).

Given that these variables pose a risk for the onset of highly comorbid disorders such as depression, anxiety disorders, insomnia, and PTSD, predict their symptom severity, contribute to their maintenance, and share common characteristics such as associations with stress, they might be regarded as transdiagnostic factors. This definition of transdiagnostic factors is consistent with criteria proposed in the literature (Dalgleish et al., 2020; Sauer-Zavala et al., 2017). The so-called mechanistically transdiagnostic constructs reveal core vulnerabilities across different mental disorders and provide insight into common mechanisms that might underly psychiatric symptoms (Harvey, Murray, Chandler, & Soehner, 2011). Even though research about transdiagnostic approaches is growing, there is a scarcity of studies that assess the mentioned variables in transdiagnostic samples. In addition, only a small number of studies have conducted such assessments by simultaneously evaluating various predictors to determine their individual, distinct effects and to control for potential interactions. Furthermore, there are few studies that have examined this topic using a transdiagnostic outcome (e.g., global symptom severity). Therefore, we aimed to address this research gap by cross-sectionally assessing the association of the following transdiagnostic factors with global symptom severity: sedentary behavior, physical activity, interaction of physical activity and physical activity-related affect regulation, repetitive negative thinking, sleep quality, perceived stress, fear of the coronavirus, emotional regulation, and emotional intelligence. Sociodemographic data such as age, gender, employment status, relationship status, and highest level of education were included as control variables. This exploration could provide crucial insights and potentially initiate the development or refinement of transdiagnostic treatment approaches that focus on those predictor variables that demonstrate a strong and significant association with global symptom severity within a sample of various mental disorders. We tested the following hypothesis: Each factor represents an individual, significant predictor of global symptom severity (i.e., transdiagnostic outcome) in a sample of German outpatients diagnosed with depression, insomnia, panic disorder, agoraphobia and/or PTSD.

## 2. Methods

### 2.1 Study design

This report is based on cross-sectional data of the baseline assessment of the ImPuls study (Wolf et al., 2024; Wolf et al., 2021). The ImPuls study was conducted according to the guidelines of the Declaration of Helsinki of 2010 and was approved by local ethics committee for medical research at the University of Tübingen (ID: 888/2020B01, 02/11/2020). The study was registered in the German Clinical Trial Register (ID: DRKS00024152, 05/02/2021). The current analysis was preregistered on Open Science Framework (https://osf.io/jhguz) before access to the cross-sectional data was distributed on March 21^st^, 2023. The preregistration was amended as it was necessary to exclude health-related quality of life from the analysis (see explanation in supplementary file S1).

### 2.2 Participants

Participants were recruited through inpatient psychiatric departments, general practitioners, psychiatric and psychotherapeutic outpatient units, (social) media and two major German health insurances [Allgemeine Ortskrankenkasse Baden-Württemberg (AOK BW) and Techniker Krankenkasse (TK)]. Participants were included if the following inclusion criteria were fulfilled: age between 18 and 65 years, fluent in German, membership of the insurance companies AOK BW or TK, no medical contraindications for exercise (participants needed to confirm their ability to exercise through a medical consultation prior to the intervention) and at least one of the following diagnoses according to ICD-10: major depressive disorders (F32.1, F32.2, F33.1, F33.2), insomnia (F51.0), panic disorder (F41.0), agoraphobia (F40.0, F40.01) and PTSD (F43.1). Exclusion criteria included acute mental and behavioral disorders due to psychotropic substances (F10.0, F10.2-F10.9; F11.0, F11.2-F11.9; F12.0, F12.2-F12.9; F13.0, F13.2-F13.9; F14.0, F14.2-F14.9; F15.0, F15.2-F15.9; F16.0, F16.2-F16.9; F17.2-F17.9; F18.0, F18.2-F18.9; F19.0, F19.2-F19.9), acute eating disorders (F50), acute bipolar disorder (F31), acute schizophrenia (F20), acute suicidality, regular engagement in at least moderate-intensity exercise for at least 30 minutes, more than once a week, continuously over a six-week period within the last three months prior to study diagnosis and medical contraindication determined by a general practitioner or a specialized medical professional.

The statistical power analysis for this report was calculated using G*Power, version 3.1.9.4 (Faul, Erdfelder, Buchner, & Lang, 2009; Faul, Erdfelder, Lang, & Buchner, 2007). With a proposed minimal effect of *f*^2^ = 0.15 (J. Cohen, 1988), number of predictors = 25, power = 0.80 and *α* = .05 the calculation resulted in a required minimum sample size of *n* = 172 for the current analysis.

Out of 1284 individuals who were screened for eligibility, 600 provided informed consent and underwent the structural diagnostic interview. Among these, 199 were excluded based on the inclusion and exclusion diagnoses, resulting in 401 individuals to be included in the cross-sectional analysis. Figure 1 illustrates the flow of participants. Baseline demographic and clinical characteristic of the sample are shown in Table 1. The mean age was 42.08 years (*SD*=13.26, range=19-65) and 71.32% identified as female. Based on the structural diagnostic interview for DSM-5, 72.1% of participants were diagnosed with depression (*n*=289), 11.5% with panic disorder (*n*=46), 9.2% with agoraphobia (*n*=37), 18.0% with PTSD (*n*=72) and 20.2% with primary insomnia (*n*=81). 98 participants (24.4%) additionally had at least one of the other inclusion diagnoses. The mean global symptom severity at baseline assessment (*M*=22.03, *SD*=11.11) was comparable to the German clinical norm sample (*M*=20.23, *SD*=12.19) (Franke, 2017).

**Figure 1.**
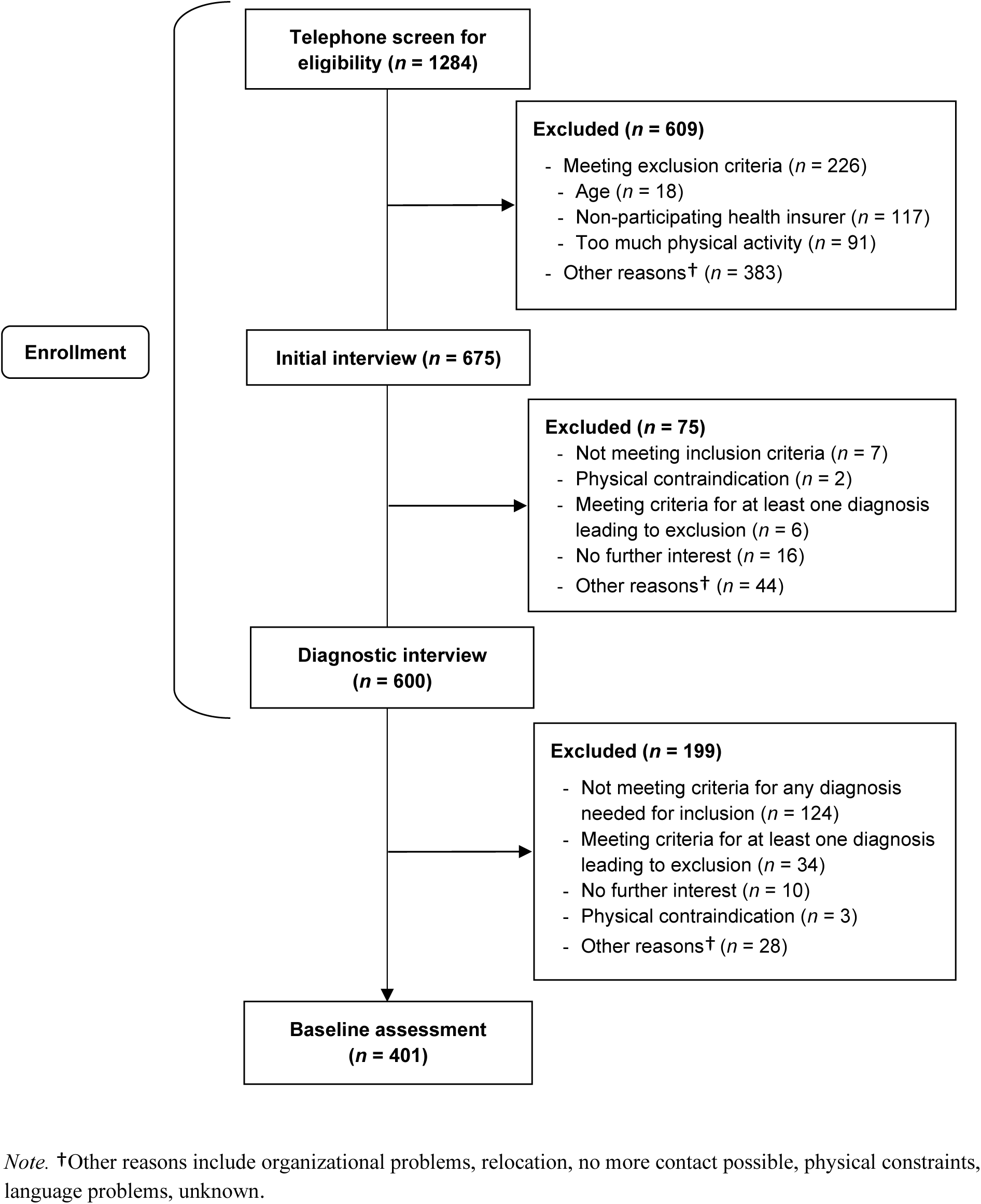
Flow of the participants up to the baseline assessment that is included in the current report.

**Table 1.**
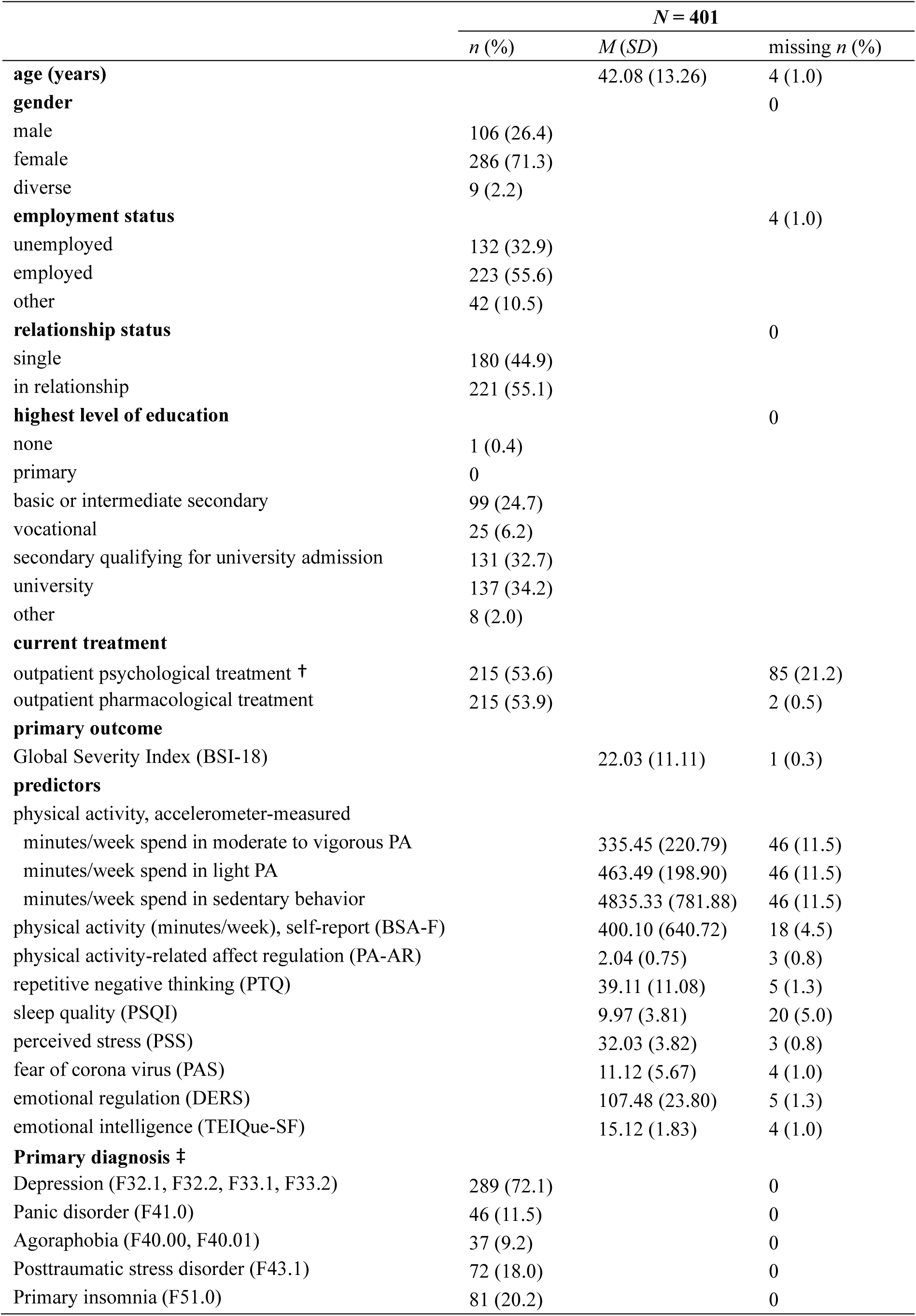
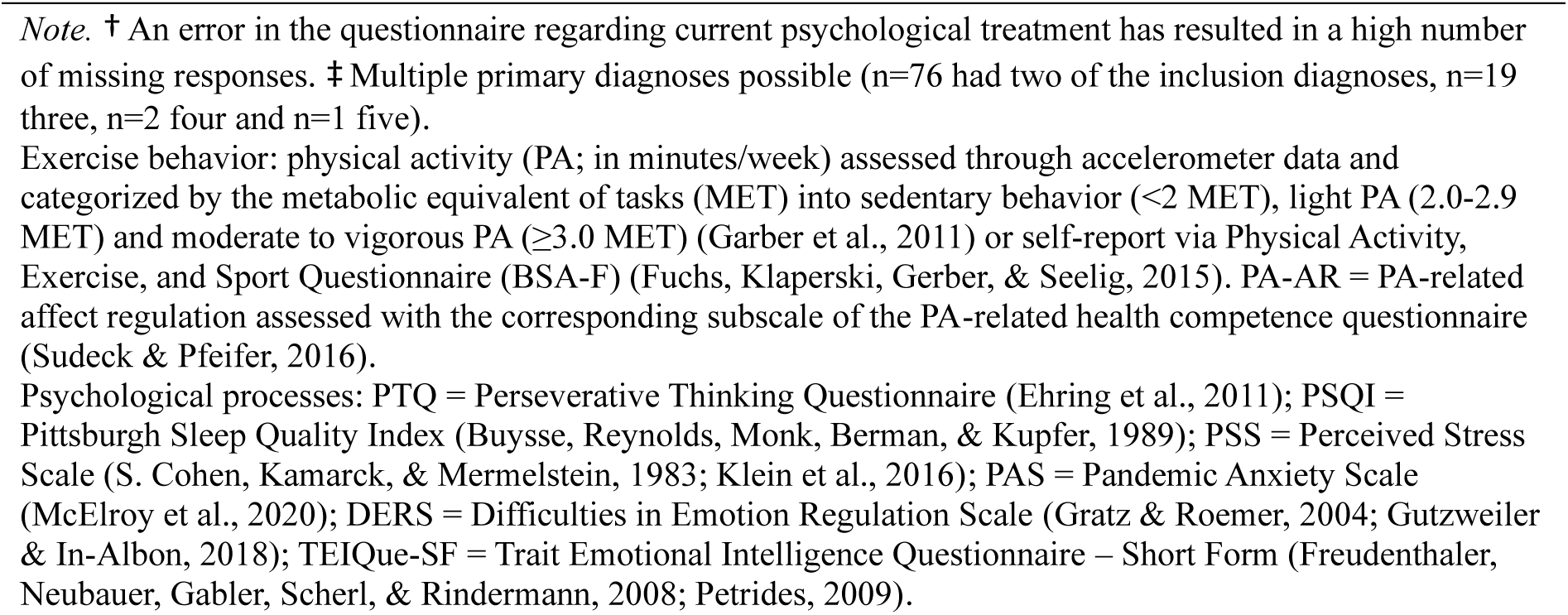
Baseline demographic and clinical characteristics of participants.

|  | *N* = 401 |  |  |
| --- | --- | --- | --- |
|  | *n* (%) | *M* (*SD*) | missing *n* (%) |
| **age (years)** |  | 42.08 (13.26) | 4 (1.0) |
| **gender** |  |  | 0 |
| male | 106 (26.4) |  |  |
| female | 286 (71.3) |  |  |
| diverse | 9 (2.2) |  |  |
| **employment status** |  |  | 4 (1.0) |
| unemployed | 132 (32.9) |  |  |
| employed | 223 (55.6) |  |  |
| other | 42 (10.5) |  |  |
| **relationship status** |  |  | 0 |
| single | 180 (44.9) |  |  |
| in relationship | 221 (55.1) |  |  |
| **highest level of education** |  |  | 0 |
| none | 1 (0.4) |  |  |
| primary | 0 |  |  |
| basic or intermediate secondary | 99 (24.7) |  |  |
| vocational | 25 (6.2) |  |  |
| secondary qualifying for university admission | 131 (32.7) |  |  |
| university | 137 (34.2) |  |  |
| other | 8 (2.0) |  |  |
| **current treatment** |  |  |  |
| outpatient psychological treatment † | 215 (53.6) |  | 85 (21.2) |
| outpatient pharmacological treatment | 215 (53.9) |  | 2 (0.5) |
| **primary outcome** |  |  |  |
| Global Severity Index (BSI-18) |  | 22.03 (11.11) | 1 (0.3) |
| **predictors** |  |  |  |
| physical activity, accelerometer-measured |  |  |  |
| minutes/week spend in moderate to vigorous PA |  | 335.45 (220.79) | 46 (11.5) |
| minutes/week spend in light PA |  | 463.49 (198.90) | 46 (11.5) |
| minutes/week spend in sedentary behavior |  | 4835.33 (781.88) | 46 (11.5) |
| physical activity (minutes/week), self-report (BSA-F) |  | 400.10 (640.72) | 18 (4.5) |
| physical activity-related affect regulation (PA-AR) |  | 2.04 (0.75) | 3 (0.8) |
| repetitive negative thinking (PTQ) |  | 39.11 (11.08) | 5 (1.3) |
| sleep quality (PSQI) |  | 9.97 (3.81) | 20 (5.0) |
| perceived stress (PSS) |  | 32.03 (3.82) | 3 (0.8) |
| fear of corona virus (PAS) |  | 11.12 (5.67) | 4 (1.0) |
| emotional regulation (DERS) |  | 107.48 (23.80) | 5 (1.3) |
| emotional intelligence (TEIQue-SF) |  | 15.12 (1.83) | 4 (1.0) |
| **Primary diagnosis ‡** |  |  |  |
| Depression (F32.1, F32.2, F33.1, F33.2) | 289 (72.1) |  | 0 |
| Panic disorder (F41.0) | 46 (11.5) |  | 0 |
| Agoraphobia (F40.00, F40.01) | 37 (9.2) |  | 0 |
| Posttraumatic stress disorder (F43.1) | 72 (18.0) |  | 0 |
| Primary insomnia (F51.0) | 81 (20.2) |  | 0 |

*Note.* † An error in the questionnaire regarding current psychological treatment has resulted in a high number of missing responses. ‡ Multiple primary diagnoses possible (n=76 had two of the inclusion diagnoses, n=19 three, n=2 four and n=1 five).
Exercise behavior: physical activity (PA; in minutes/week) assessed through accelerometer data and categorized by the metabolic equivalent of tasks (MET) into sedentary behavior (<2 MET), light PA (2.0-2.9 MET) and moderate to vigorous PA ( $\geq 3.0$ MET) (Garber et al., 2011) or self-report via Physical Activity, Exercise, and Sport Questionnaire (BSA-F) (Fuchs, Klaperski, Gerber, & Seelig, 2015). PA-AR = PA-related affect regulation assessed with the corresponding subscale of the PA-related health competence questionnaire (Sudeck & Pfeifer, 2016).
Psychological processes: PTQ = Perseverative Thinking Questionnaire (Ehring et al., 2011); PSQI = Pittsburgh Sleep Quality Index (Buysse, Reynolds, Monk, Berman, & Kupfer, 1989); PSS = Perceived Stress Scale (S. Cohen, Kamarck, & Mermelstein, 1983; Klein et al., 2016); PAS = Pandemic Anxiety Scale (McElroy et al., 2020); DERS = Difficulties in Emotion Regulation Scale (Gratz & Roemer, 2004; Gutzweiler & In-Albon, 2018); TEIQue-SF = Trait Emotional Intelligence Questionnaire – Short Form (Freudenthaler, Neubauer, Gabler, Scherl, & Rindermann, 2008; Petrides, 2009).

### 2.3 Procedure

Data was collected at 10 different study sites in Baden-Württemberg, Germany. The active enrolment period lasted from March 2021 until May 2022. Interested participants first attended a preliminary phone contact where they received general project information and completed screenings of eligibility criteria and somatic contraindications for exercise. Potentially eligible participants were then invited to an inhouse meeting taking place in the study site closest to their residence. They were informed about study procedures, provided written informed consent and were screened initially for exclusion diagnoses to prepare for the following structural diagnostic interview for DSM-5 (SCID-5-CV) (Beesdo-Baum, Zaudig, & Wittchen, 2019) with a trained study psychologist. Once six participants at one study site were eligible for participation, they received online questionnaires via the web-based data management system REDCap (P. A. Harris et al., 2019; P. A. Harris et al., 2009) which could be answered within a 14-days-period. Additionally, they wore accelerometer-based physical activity sensors (MOVE 4; movisens GmbH) for seven consecutive days.

### 2.4 Outcomes

#### 2.4.1 Primary outcome

Global symptom severity served as the primary outcome and was measured using the Global Severity Index (GSI), derived from the German version of the Brief Symptom Inventory-18 [BSI-18] (Franke, 2017). The GSI encompasses ratings of general mental distress across somatization, depression, and anxiety symptom scales. Each symptom scale consists of six items, resulting in a total of 18 items that were rated on a 5-point Likert scale (range: 0-4). The total score for each scale is calculated, and the GSI is obtained by summing these three scores. Higher scores on the GSI indicate greater levels of distress, with a clinical cut-off set at 12. Among patients with various mental disorders, the GSI has demonstrated good internal consistency (*α*=.89) (Franke, 2017). Cronbach’s alpha of the GSI at baseline assessment in our study was *α*=.86, indicating good internal consistency.

#### 2.4.2 Predictor and control variables

Accelerometer-measured and self-reported physical activity, sedentary behavior, the interaction of accelerometer-measured and self-reported physical activity and physical activity-related affect regulation, repetitive negative thinking, sleep quality, perceived stress, fear of the coronavirus, emotional regulation, and emotional intelligence served as predictors, Sociodemographic data (i.e., age, gender, employment status, relationship status, highest level of education) were included as control variables. The measurement instruments (German versions) used are shown in Table 2. A detailed description of the predictor and control variables and the corresponding questionnaires is in supplementary file S2.

**Table 2.**
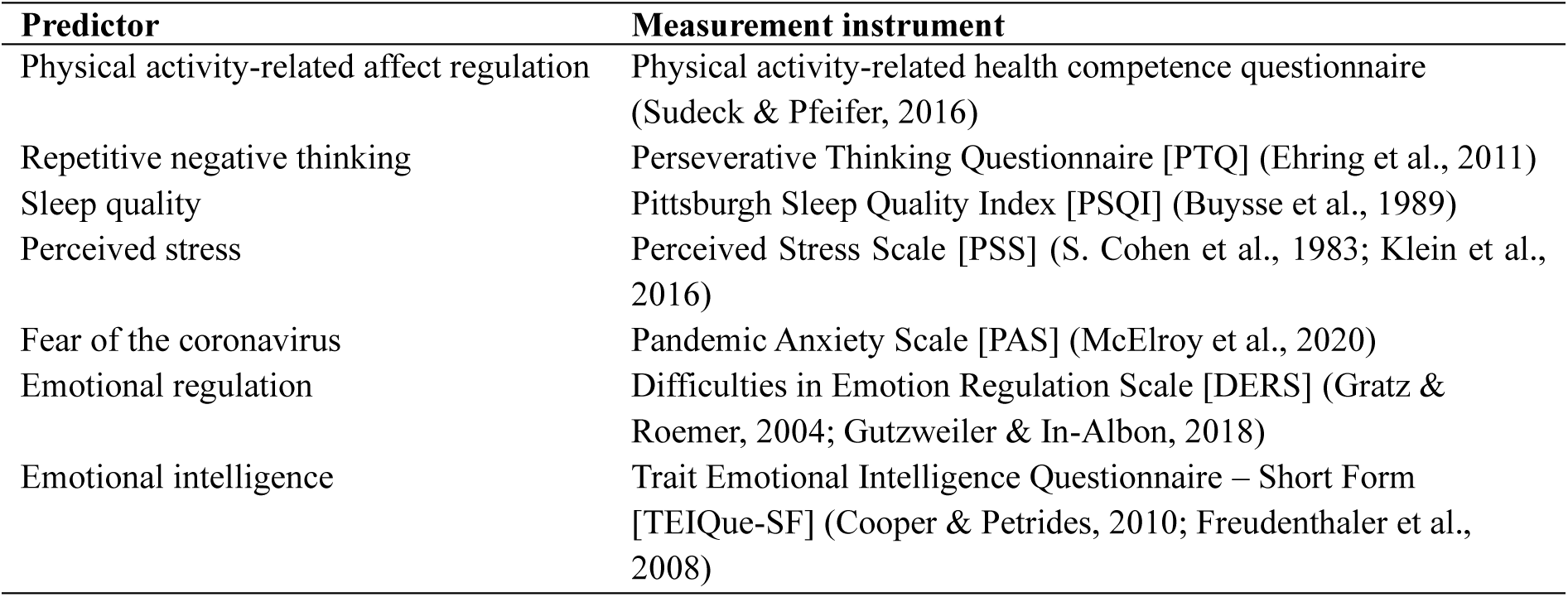
Measurement instruments used to assess the predictor variables.

| Predictor | Measurement instrument |
| --- | --- |
| Physical activity-related affect regulation | Physical activity-related health competence questionnaire (Sudeck & Pfeifer, 2016) |
| Repetitive negative thinking | Perseverative Thinking Questionnaire [PTQ] (Ehring et al., 2011) |
| Sleep quality | Pittsburgh Sleep Quality Index [PSQI] (Buysse et al., 1989) |
| Perceived stress | Perceived Stress Scale [PSS] (S. Cohen et al., 1983; Klein et al., 2016) |
| Fear of the coronavirus | Pandemic Anxiety Scale [PAS] (McElroy et al., 2020) |
| Emotional regulation | Difficulties in Emotion Regulation Scale [DERS] (Gratz & Roemer, 2004; Gutzweiler & In-Albon, 2018) |
| Emotional intelligence | Trait Emotional Intelligence Questionnaire – Short Form [TEIQue-SF] (Cooper & Petrides, 2010; Freudenthaler et al., 2008) |

### 2.5 Statistical analysis

Data preparation and analysis were carried out using R, version 4.3.3 (R Core Team, 2024). The analytic code is available at https://osf.io/5rcuz/files/osfstorage.

Descriptive statistics were used to analyze sample characteristics and are reported in frequencies (*n*) and percentages (%) for categorical variables and in means (*M*) and standard deviations (*SD*) for continuous variables. Self-reported physical activity was manually checked for plausibility for values greater than 2000 minutes per week for the Physical Activity Index as well as greater than 200 minutes per week for the Exercise Index. Altogether, eleven values were omitted because they appeared unrealistic: ten from the Physical Activity Index and one from the Exercise Index. Accelerometer-measured data was included if at least four consecutive days with at least eight hours of wearing time were recorded as recommended by current guidelines (Donaldson, Montoye, Tuttle, & Kaminsky, 2016; Migueles et al., 2017). We computed a mean activity value of the valid days and multiplied the result by seven to obtain a measure of minutes per week. All values were visually checked for normality of distribution. Accelerometer-based moderate to vigorous physical activity, self-reported physical activity and physical activity-related affect regulation were log-transformed due to skewness of data.

Potential outliers of all variables included in the analysis were identified through three measures: interquartile range, Leverage and Cook’s Distance. Thus, data points that are more than 1.5 times the interquartile range away from the 25th and 75th percentile, with a leverage of greater than 0.19 (J. Cohen, Cohen, West, & Aiken, 2002), and a Cook’s distance of greater than 0.5, were considered as potential outliers. A total of 114 participants were excluded according to these proposed measures. The number of exclusions can be attributed to skewness of data and the presence of several extreme predictor values. Multiple linear regression was calculated again as a sensitivity analysis to evaluate the influence of potential outliers.

Data analysis of the primary outcome was performed using multiple linear regression. The research question was analyzed with accelerometer-measured and self-reported physical activity, sedentary behavior, the interaction of accelerometer-measured and self-reported physical activity and physical activity-related affect regulation as well as repetitive negative thinking, sleep quality, perceived stress, fear of the coronavirus, emotional regulation, and emotional intelligence as predictors and global symptom severity as the outcome. Sociodemographic data were included as control variables. Statistical significance was defined as a *p* value *<*.05. Assumptions of multiple linear regression (i.e., linearity, normality of the residuals, homoscedasticity, and multicollinearity) were visually inspected and verified by statistical tests (i.e., Rainbow test for linearity, Kolmogorov-Smirnov test for normality of the residuals, Breusch-Pagan test for homoscedasticity, Variance Inflation Factor for multicollinearity). Standardized regression coefficients were calculated.

## 3. Results

### 3.1 Missing values

After data collection, we identified 2.8% of missing data at scale level with 372 of 401 participants having at least one missing (92.8%). We used (scale-based) multiple imputation to create 10 imputed datasets that contained all variables included in the current report. All analyses were then conducted using the 10 imputed datasets and results were pooled according to Rubin’ rules (Rubin, 1987).

### 3.2 Multiple regression analysis

The model including accelerometer-measured and self-reported physical activity, sedentary behavior, the interaction of accelerometer-measured and self-reported physical activity and physical activity-related affect regulation, repetitive negative thinking, sleep quality, perceived stress, fear of the coronavirus, emotional regulation, and emotional intelligence as independent variables (predictors), sociodemographic data as control variables and global symptom severity as the dependent variable (outcome) explained 40.3% of variance on the GSI (adjusted *R^2^*=0.403). Emotional regulation (β=.256, *p*<.001), sleep quality (β=.221, *p*<.001), perceived stress (β=.217, *p*<.001), repetitive negative thinking (β=.214, *p*<.001), emotional intelligence (β=-.090, *p*=.034) and fear of the coronavirus (β=.088, *p*=.036) were significantly associated with global symptom severity. There were no significant associations between either sociodemographic data, physical activity, or sedentary behavior and global symptom severity. The full statistics and regression coefficients are presented in Table 3. The Pearson’s correlation matrix of global symptom severity and predictor variables are provided in supplementary file S3.

**Table 3.**
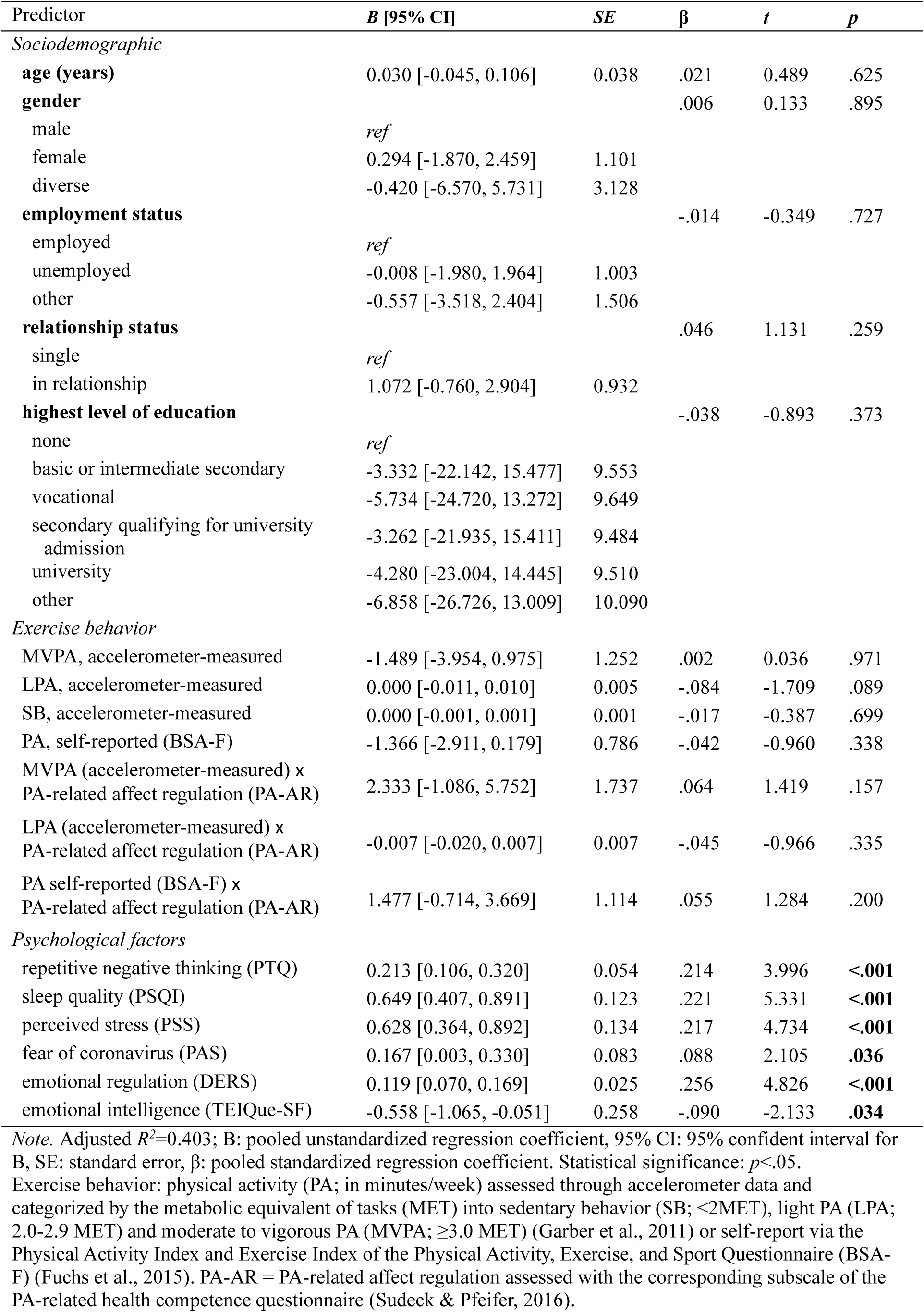
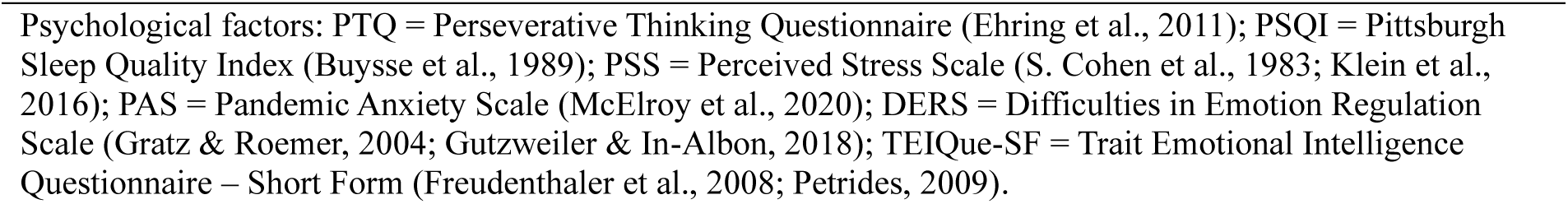
Multiple regression for global symptom severity (*N* = 401)

| Predictor | *B* [95% CI] | *SE* | $\beta$ | *t* | *p* |
| --- | --- | --- | --- | --- | --- |
| *Sociodemographic* |  |  |  |  |  |
| **age (years)** | 0.030 [-0.045, 0.106] | 0.038 | .021 | 0.489 | .625 |
| **gender** |  |  | .006 | 0.133 | .895 |
| male | *ref* |  |  |  |  |
| female | 0.294 [-1.870, 2.459] | 1.101 |  |  |  |
| diverse | -0.420 [-6.570, 5.731] | 3.128 |  |  |  |
| **employment status** |  |  | -.014 | -0.349 | .727 |
| employed | *ref* |  |  |  |  |
| unemployed | -0.008 [-1.980, 1.964] | 1.003 |  |  |  |
| other | -0.557 [-3.518, 2.404] | 1.506 |  |  |  |
| **relationship status** |  |  | .046 | 1.131 | .259 |
| single | *ref* |  |  |  |  |
| in relationship | 1.072 [-0.760, 2.904] | 0.932 |  |  |  |
| **highest level of education** |  |  | -.038 | -0.893 | .373 |
| none | *ref* |  |  |  |  |
| basic or intermediate secondary | -3.332 [-22.142, 15.477] | 9.553 |  |  |  |
| vocational | -5.734 [-24.720, 13.272] | 9.649 |  |  |  |
| secondary qualifying for university admission | -3.262 [-21.935, 15.411] | 9.484 |  |  |  |
| university | -4.280 [-23.004, 14.445] | 9.510 |  |  |  |
| other | -6.858 [-26.726, 13.009] | 10.090 |  |  |  |
| *Exercise behavior* |  |  |  |  |  |
| MVPA, accelerometer-measured | -1.489 [-3.954, 0.975] | 1.252 | .002 | 0.036 | .971 |
| LPA, accelerometer-measured | 0.000 [-0.011, 0.010] | 0.005 | -.084 | -1.709 | .089 |
| SB, accelerometer-measured | 0.000 [-0.001, 0.001] | 0.001 | -.017 | -0.387 | .699 |
| PA, self-reported (BSA-F) | -1.366 [-2.911, 0.179] | 0.786 | -.042 | -0.960 | .338 |
| MVPA (accelerometer-measured) x PA-related affect regulation (PA-AR) | 2.333 [-1.086, 5.752] | 1.737 | .064 | 1.419 | .157 |
| LPA (accelerometer-measured) x PA-related affect regulation (PA-AR) | -0.007 [-0.020, 0.007] | 0.007 | -.045 | -0.966 | .335 |
| PA self-reported (BSA-F) x PA-related affect regulation (PA-AR) | 1.477 [-0.714, 3.669] | 1.114 | .055 | 1.284 | .200 |
| *Psychological factors* |  |  |  |  |  |
| repetitive negative thinking (PTQ) | 0.213 [0.106, 0.320] | 0.054 | .214 | 3.996 | <.001 |
| sleep quality (PSQI) | 0.649 [0.407, 0.891] | 0.123 | .221 | 5.331 | <.001 |
| perceived stress (PSS) | 0.628 [0.364, 0.892] | 0.134 | .217 | 4.734 | <.001 |
| fear of coronavirus (PAS) | 0.167 [0.003, 0.330] | 0.083 | .088 | 2.105 | .036 |
| emotional regulation (DERS) | 0.119 [0.070, 0.169] | 0.025 | .256 | 4.826 | <.001 |
| emotional intelligence (TEIQue-SF) | -0.558 [-1.065, -0.051] | 0.258 | -.090 | -2.133 | .034 |

*Note.* Adjusted $R^2=0.403$ ; *B*: pooled unstandardized regression coefficient, 95% CI: 95% confident interval for *B*, *SE*: standard error, $\beta$ : pooled standardized regression coefficient. Statistical significance: $p<.05$ .
Exercise behavior: physical activity (PA; in minutes/week) assessed through accelerometer data and categorized by the metabolic equivalent of tasks (MET) into sedentary behavior (SB; <2MET), light PA (LPA; 2.0-2.9 MET) and moderate to vigorous PA (MVPA; $\geq 3.0$ MET) (Garber et al., 2011) or self-report via the Physical Activity Index and Exercise Index of the Physical Activity, Exercise, and Sport Questionnaire (BSA-F) (Fuchs et al., 2015). PA-AR = PA-related affect regulation assessed with the corresponding subscale of the PA-related health competence questionnaire (Sudeck & Pfeifer, 2016).
Psychological factors: PTQ = Perseverative Thinking Questionnaire (Ehring et al., 2011); PSQI = Pittsburgh Sleep Quality Index (Buysse et al., 1989); PSS = Perceived Stress Scale (S. Cohen et al., 1983; Klein et al., 2016); PAS = Pandemic Anxiety Scale (McElroy et al., 2020); DERS = Difficulties in Emotion Regulation Scale (Gratz & Roemer, 2004; Gutzweiler & In-Albon, 2018); TEIQue-SF = Trait Emotional Intelligence Questionnaire – Short Form (Freudenthaler et al., 2008; Petrides, 2009).

### 3.3 Sensitivity analysis

The sensitivity analysis after exclusion of identified outliers revealed comparable results except for fear of the coronavirus and emotional intelligence, which were no longer significant (fear of the coronavirus: β=.093, *p*=.072; emotional intelligence: β=-.052, *p*=.314). The detailed results of the sensitivity analysis including statistics and regression coefficients are provided in supplementary file S4.

## 4. Discussion

In this study, we examined cross-sectional associations of transdiagnostic predictors of global symptom severity among outpatients diagnosed with depressive disorders, non-organic primary insomnia, agoraphobia, panic disorder, and/or PTSD. Higher levels of repetitive negative thinking, fear of the coronavirus and perceived stress, worse sleep quality as well as greater difficulties with emotional regulation and emotional intelligence – thus, transdiagnostic psychological processes typically involved in the adaption to stressful life events – were distinctly, significantly associated with higher global symptom severity in our outpatient sample when controlling for sociodemographic data and all other predictors in a multiple regression analysis. However, contrary to our hypothesis, no associations between global symptom severity and the amount of daily physical activity and sedentary behavior were found. The bivariate correlations showed consistent results between the predictors and global symptom severity, except for emotional intelligence, which was not significantly correlated with global symptom severity. The sensitivity analysis after exclusion of identified outliers also revealed comparable results. Hence, the results suggest that repetitive negative thinking, perceived stress, sleep quality, and emotional regulation represent the most important transdiagnostic factors in our transdiagnostic sample. Interestingly, repetitive negative thinking and emotional regulation demonstrated significant correlations with each other and with perceived stress, highlighting their interconnectedness in the context of stress.

Importantly, literature suggests that each psychological factor plays a major role in the onset and maintenance of the included mental disorders and/or contributes to their severity of symptoms. There is evidence that some of the examined factors are associated with several clinically meaningful consequences. Heightened levels of repetitive negative thinking, sleep disturbances, experiencing stressful life events, and difficulties with emotion regulation, assessed in clinical and general populations, were associated with increased risk of suicide behavior, mainly suicide ideation (Caudle et al., 2024; Colmenero-Navarrete, Garcia-Sancho, & Salguero, 2022; L. M. Harris, Huang, Linthicum, Bryen, & Ribeiro, 2020; Howarth et al., 2020). In addition, among patients receiving treatment for mental disorders such as depression and anxiety disorders, sleep disturbances were associated with diminished quality of life and global functioning (Kallestad et al., 2012). Likewise, feelings of perceived distress and a sense of lack of control appeared to be related to impaired quality of life in individuals with mood and stress-related disorders (Connell, Brazier, O’Cathain, Lloyd-Jones, & Paisley, 2012).

Importantly, up to this point, these psychological factors have primarily been assessed in relation to specific mental disorders (Scott, Webb, Martyn-St James, Rowse, & Weich, 2021; Sloan et al., 2017; Wahl et al., 2019; Zorn et al., 2017). The findings of this study therefore represent an important expansion to existing literature by demonstrating that underlying mechanisms predominantly investigated in disorder-specific samples also hold relevance in transdiagnostic contexts. The results suggest that repetitive negative thinking, emotional regulation, sleep quality, and the experience of stress might be core transdiagnostic factors that are present across various stress-related mental disorders. From a theoretical perspective, these findings lend further support to transdiagnostic approaches and the potential clinical relevance lies in the identification of psychological factors that could serve as targets for transdiagnostic interventions, specifically for stress-associated disorders. The results are particularly relevant given the limited number of studies exploring psychological predictors of transdiagnostic outcomes such as global symptom severity within transdiagnostic outpatient samples.

Another noteworthy point is that the examined psychological factors had a distinct, significant association with global symptom severity, even though some were significantly correlated with each other and share common characteristics, such as an association with stress. Therefore, this study is an important extension to previous research by emphasizing that these psychological factors are theoretically interrelated and correlate with each other yet maintain distinct and individual associations with global symptom severity in transdiagnostic samples. This finding indicates the presence of distinct processes and unique vulnerability factors.

Given their high prevalence and clinical significance it is expected that these psychological factors might be prime targets of interventions. Indeed, strategies aimed at reducing repetitive negative thinking, enhancing stress management skills, improving sleep hygiene, and fostering emotional regulation are core components of many disorder-specific psychological interventions. Our study results suggest that these factors could also be considered as transdiagnostic treatment targets. Consequently, the current study may serve as a crucial foundation for the development of evidence-based transdiagnostic interventions.

Contrary to our hypothesis, no significant associations were found for either daily physical activity or sedentary behavior and global symptom severity. This report is based on cross-sectional data of the baseline assessment of an exercise intervention trial aimed at assessing the therapeutic impact of exercise. Therefore, only individuals with low physical activity levels who did not engage in exercise for more than 30 minutes per week in the last three months before the study diagnosis were included. This specific inclusion criterion minimized variance of the physical activity variable within the sample and the resulting loss of information might have led to the non-significant association with global symptom severity. Additionally, the amount of physical activity analyzed in this study primarily captured routine daily activities such as transportation, household chores, leisure-time pursuits, and occupational tasks. So far, only few studies have explored the impact of these everyday physical activities on mental health. Whereas leisure-time physical activity is related with positive mental health outcomes, meta-analytic findings suggest inconsistent associations with the other domains (White et al., 2017). In fact, there is evidence indicating that physical activity performed in the domains of occupational, transportation, and domestic was associated with increased depressive symptoms (Lopes et al., 2023) and housework was also related with more experienced psychological distress (Asztalos et al., 2009). One possible explanation for these findings includes the obligatory nature of these activities. Household chores are necessary tasks rarely chosen for enjoyment. Similarly, while e.g., biking to work might generally be a pleasurable activity, its purpose – commuting – makes it a necessity and therefore less enjoyable. Consequently, these activities are unlikely to be chosen to specifically regulate affect, which may account for the lack of interaction effects in our analysis.

Sociodemographic data were not significantly associated with global symptom severity over and above psychological variables in our model. This finding suggests that in an outpatient sample already diagnosed with and suffering from mental disorders, primarily psychological variables account for variance on global symptom severity. Sociodemographic factors such as being female, a young adult, single, unemployed or retired, and belonging to lower social class seem to increase the likelihood of becoming prevalent with a mental disorder (Jacobi et al., 2014; Jacobi et al., 2004) rather than explaining variance of psychopathology within a sample of mentally ill patients. Thus, these sociodemographic factors are considered more distal and do not directly influence psychopathology; instead, their effects are mediated by more proximal processes, such as psychological factors. If the selection of proximal processes is sufficiently comprehensive, no direct effects of distal variables (i.e., sociodemographic data) are to be expected in this context. In addition, the finding that non-modifiable sociodemographic variables show no significant association with global symptom severity, in contrast to modifiable psychological variables which do, and account for a significant amount of variance, supports the idea of reducing global symptom severity in patients through transdiagnostic interventions.

It is important to recognize that the actual sample size was more than twice as large as the minimum sample size required, determined by power calculation. A post hoc power calculation with the given sample size of 401 revealed a power of 99.9% for the current report (Faul et al., 2009; Faul et al., 2007). However, when interpreting results of overpowered studies, statistical significance should not be confused with clinical meaningful effects (Sedgwick, 2013). Therefore, it is crucial to consider the reported effect sizes (β) and the degree of variance explanation (adjusted R^2^), alongside mere p-values. Effect sizes of β=.256 for emotional regulation, β=.221 for sleep quality, β=.217 for perceived stress, and β=.214 for repetitive negative thinking as well as an explained variance of 40.3% on global symptom severity can be regarded acceptable in social science research given the complexity of human behavior and the number of potential unmeasured factors. These results become even more convincing when considering that the majority of our sample received psychological (68%) or pharmacological (54%) treatment at the time of assessment (see Table 2) and still, the psychological factors were strongly and significantly associated with global symptom severity.

### 4.1 Strengths and limitations

A key strength of this study comprises the large transdiagnostic sample consisting of German outpatients whose clinical diagnoses were verified through structured interviews conducted by trained study psychologists. The study sample further enables the generalization of results to the German population diagnosed with mental disorders. The mean age of our sample, 42.1 years, aligns with the mean age of the general population, which is 44.6 years (Federal Statistical Office, 2023). Additionally, the overrepresentation of female participants corresponds with the higher incidence of mental disorders, such as affective and anxiety disorders, among women. Notably, depression is considered more prevalent than the other diagnoses included in the study (Jacobi et al., 2014, 2016). Furthermore, all predictors were measured by validated and widely used scales or accelerometer-based sensors and an external institution was responsible for data management, including data collection, storing and anonymization. In addition, participants were blinded to the research question, which minimized potential biases and a wide range of potential predictors of global symptom severity were simultaneously investigated within a transdiagnostic sample. The study also faces several limitations as the cross-sectional research design which makes it impossible to draw causal conclusions. It is crucial to conduct additional prospective and longitudinal studies for a more thorough exploration of the causal relationships involved. In addition, the sample was restricted as only physically inactive individuals were included, i.e., participants who had exercised continuously for at least 30 minutes at least twice a week in the last three months before the study diagnosis were excluded. This criterion particularly limited leisure-time physical activity, which is the domain typically associated with mental health. Excluding physically active individuals restricts the generalizability of our results regarding physical activity and may have contributed to the lack of significant associations with global symptom severity.

### 4.2 Conclusion and future directions

This report examined cross-sectional associations of several variables (i.e., psychological factors, physical activity, sedentary behavior) with global symptom severity among outpatients with various stress-related mental disorders. The findings highlight that common and prevalent psychological processes as repetitive negative thinking, perceived stress, sleep quality, and emotional regulation, in contrast to physical activity, and sedentary behavior, each have a significant association with global transdiagnostic symptom severity among patients diagnosed with depressive disorders, non-organic primary insomnia, agoraphobia, panic disorder, and/or PTSD. Thus, variables predominantly studied in disorder-specific samples also hold relevance in transdiagnostic contexts. Consequently, these psychological processes can serve as targets not only in disorder-specific interventions but also in transdiagnostic treatment approaches. However, further research in prospective longitudinal studies with transdiagnostic samples is warranted.

## Data Availability

All data produced are available online at https://osf.io/5rcuz/files/osfstorage.

## Acknowledgments

The German Innovation Fund of the Federal Joint Committee of Germany fully funds the study from September 2020 to June 2024 (grant number: 01NVF19022). We thank all general practitioners, psychiatrists, psychotherapists, clinics, hospitals, social media influencers, and newspapers that supported the recruitment process. We would especially like to thank all student assistants who supported us in the recruitment.

## Data Availability Statement

The data that support the findings of this study are openly available in Open Science Framework at https://osf.io/5rcuz/files/osfstorage.

