## Supplemental data for "Associations between transdiagnostic psychological processes and global symptom severity among outpatients with various mental disorders: A cross-sectional study"

**S1.** Deviation from preregistration

We excluded health-related quality of life from our analysis as this variable does not align with our definition of an impact factor. In addition, a correlation analysis of the EQ-5D-5L questionnaire (Herdman et al., 2011; Hinz, Kohlmann, Stobel-Richter, Zenger, & Brahler, 2014) with the primary outcome (i.e., global symptom severity measured using the Global Severity Index of the Brief Symptom Inventory [BSI-18] (Franke, 2017)) demonstrated a high correlation ($r$=-.608). This strong correlation may be attributed to the fact that the domain "anxiety or depression" of the EQ-5D-5L questionnaire corresponds to the depression and anxiety subscales of the BSI-18 (Franke, 2017).

**S2.** Detailed description of predictor and control variables included in the multiple linear regression analysis

**Accelerometer-measured and self-reported physical activity (PA).** Accelerometer-measured PA was assessed via accelerometer-based sensors (Move 4, movisens GmbH). The sensor assesses PA based on kinematic data in three dimensions and atmospheric air pressure. This allows to estimate the amount of PA through step counts and of different intensities for a specified time period based on validated algorithms (Anastasopoulou, Tansella, Stumpp, Shammas, & Hey, 2012). Participants wore the sensors for seven consecutive days on the right hip. PA was defined as any activity that exceeds 1.99 metabolic equivalents of tasks (MET), with light intensity PA (LPA) requiring 2.0–2.9 METs and moderate to vigorous PA (MVPA) ≥3.0 METs. Accordingly, sedentary behavior (SB) was defined as any activity less than 2.0 MET (Garber et al., 2011). Data was included as average time spent in SB, LPA and MVPA in minutes per day. Self-reported PA in minutes per week was assessed using the self-report Physical Activity Index (including job transportation, walking, cycling, physically demanding care and housework) and the Exercise Index (including physical activity with the goal of improving fitness) of the Physical Activity, Exercise, and Sport Questionnaire (BSA questionnaire; Fuchs, Klaperski, Gerber, & Seelig, 2015). Participants specified type, duration, and frequency of PA and exercise in the last four weeks. The scores were combined to a total score of PA in minutes per week.

**PA-related affect regulation.** PA-related affect regulation was assessed with the corresponding subscale of the German questionnaire PA-related health competence (Sudeck & Pfeifer, 2016). The subscale consists of 4 items rated on a 4-point Likert scale (range: 1-4). The total score was calculated by averaging over all items with higher scores indicating greater PA-related affect regulation.

**Repetitive negative thinking.** Repetitive negative thinking was assessed with the German version of the Perseverative Thinking Questionnaire [PTQ] (Ehring et al., 2011). The PTQ consist of 15 items evaluating the assumed process characteristics of repetitive negative thinking with 3 items each (repetitive, intrusive, difficult to disengage from, unproductive, capturing mental capacity). The items are rated on a 5-point Likert scale (range: 0-4) and higher scores indicate greater repetitive negative thinking.

**Sleep Quality.** Sleep quality was assessed with the global sleep quality score of the German version of the Pittsburgh Sleep Quality Index [PSQI] (Buysse, Reynolds, Monk, Berman, & Kupfer, 1989). The global sleep quality score is the sum of seven sleep component scores (range: 0-3), including subjective sleep quality, sleep latency, sleep duration, habitual sleep efficiency, sleep disturbances, use of sleeping medications, and daytime dysfunction. Higher scores indicate worse sleep quality.

**Perceived stress.** Perceived stress was assessed with the German version of the Perceived Stress Scale [PSS] (Cohen, Kamarck, & Mermelstein, 1983; Klein et al., 2016). Ten items measure the degree to which life in the past month has been experienced as unpredictable, uncontrollable and overwhelming on a 5-point Likert scale (range: 1-5). Higher scores indicate greater perceived stress.

**Fear of the corona virus.** Fear of the corona virus was assessed with the German version of the Pandemic Anxiety Scale [PAS] (McElroy et al., 2020). The PAS exists of seven items forming a total score and assessing the subscales Disease Anxiety and Consequence Anxiety. The item scores range from 0 to 4 and higher scores indicate greater pandemic anxiety.

**Emotional regulation.** Emotional regulation was assessed with the German version of the Difficulties in Emotion Regulation Scale [DERS] (Gratz & Roemer, 2004; Gutzweiler & In-

Albon, 2018) that consists of 36 items. The total score is calculated by adding the scores of the items (range: 1-5). Higher scores indicate greater problems with emotional regulation.

**Emotional intelligence.** Emotional intelligence was assessed with the German version of the Trait Emotional Intelligence Questionnaire – Short Form [TEIQue-SF] (Cooper & Petrides, 2010; Freudenthaler, Neubauer, Gabler, Scherl, & Rindermann, 2008; Petrides, 2009). The TEIQue-SF consists of 30 items that measure 15 facets of emotional intelligence and are rated on a 7-point Likert scale (range: 1-7). Higher scores indicate greater emotional intelligence.

**Sociodemographic data.** Sociodemographic data as age, gender (i.e., female, male, diverse), employment status (i.e., employed, unemployed, other), relationship status (i.e., single, in relationship) and highest level of education (i.e., none, primary, secondary, vocational, high school diploma, university, other) were assessed via self-report as part of the demographic questionnaire administered at the same measurement as the primary outcome and the other predictors.

**S3.** Pearson's correlation matrix of global symptom severity and potential predictors

| **Variables** | 1 | 2 | 3 | 4 | 5 | 6 | 7 | 8 | 9 | 10 | 11 |
|---|---|---|---|---|---|---|---|---|---|---|---|
| 1. Global symptom severity | 1 | | | | | | | | | | |
| 2. MVPA | -0.06 | 1 | | | | | | | | | |
| 3. LPA | -0.05 | 0.44*** | 1 | | | | | | | | |
| 4. SB | -0.04 | -0.35*** | -0.27*** | 1 | | | | | | | |
| 5. PA | 0.03 | 0.08 | 0.18* | 0.01 | 1 | | | | | | |
| 6. repetitive negative thinking | 0.48*** | 0.10 | 0.05 | -0.12 | -0.01 | 1 | | | | | |
| 7. sleep quality | 0.34*** | -0.10 | 0.05 | -0.01 | 0.09 | 0.12 | 1 | | | | |
| 8. perceived stress | 0.39*** | -0.01 | 0.10 | -0.08 | 0.02 | 0.39*** | 0.15 | 1 | | | |
| 9. fear of coronavirus | 0.18* | -0.07 | 0.00 | -0.01 | 0.08 | 0.10 | 0.09 | 0.22*** | 1 | | |
| 10. emotional regulation | 0.49*** | -0.01 | -0.07 | -0.03 | -0.10 | 0.63*** | 0.16 | 0.27*** | 0.07 | 1 | |
| 11. emotional intelligence | 0.00 | -0.01 | -0.01 | -0.05 | -0.01 | 0.13 | -0.05 | 0.20** | 0.14 | 0.05 | 1 |

*Note.* MVPA=moderate to vigorous physical activity, accelerometer-measured; LPA=light physical activity, accelerometer-measured; SB=sedentary behavior, accelerometer-measured; PA=physical activity, self-reported.

For reasons of clarity, the bivariate correlations with the control variables (i.e., sociodemographic data) were excluded from this correlation matrix. Significant correlations were found for the following variables: Age was significantly correlated with LPA ($r$=0.19*), repetitive negative thinking ($r$=-0.18*), and emotional regulation ($r$=-0.23***).

***$p$<.001, **$p$<.01, *$p$<.05.

**S4.** Multiple regression for GSI after exclusion of identified outliers ($N = 287$)

| Predictor | ***B* [95% CI]** | ***SE*** | **β** | ***t*** | ***p*** |
|---|---|---|---|---|---|
| *Sociodemographic* | | | | | |
| **age (years)** | -0.038 [-0.128, 0.051] | 0.091 | -.063 | -1.143 | .254 |
| **gender** | | | .050 | 0.949 | .348 |
| male | *ref* | | | | |
| female | 1.215 [-1.334, 3.764] | 2.588 | | | |
| diverse | 1.756 [-9.085, 12.596] | 10.991 | | | |
| **employment status** | | | .007 | 0.137 | .891 |
| employed | *ref* | | | | |
| unemployed | 0.337 [-2.055, 2.729] | 2.429 | | | |
| other | -0.049 [-3.613, 3.515] | 3.618 | | | |
| **relationship status** | | | .091 | 1.785 | .075 |
| single | *ref* | | | | |
| in relationship | 2.038 [-0.123, 4.198] | 2.194 | | | |
| **highest level of education** | | | -.030 | -0.548 | .584 |
| basic or intermediate secondary | *ref* | | | | |
| vocational | -4.047 [-9.299, 1.204] | 5.334 | | | |
| secondary qualifying for university admission | 0.133 [-2.871, 3.137] | 3.050 | | | |
| university | -0.958 [-3.946, 2.029] | 3.034 | | | |
| other | -7.090 [-19.800, 5.621] | 12.906 | | | |
| *Exercise behavior* | | | | | |
| MVPA, accelerometer-measured | -2.043 [-5.331, 1.244] | 3.333 | -.003 | -0.044 | .965 |
| LPA, accelerometer-measured | 0.000 [-0.015, 0.014] | 0.015 | -.045 | -0.718 | .474 |
| SB, accelerometer-measured | 0.000 [-0.002, 0.002] | 0.002 | .028 | 0.477 | .634 |
| PA, self-reported (BSA-F) | -1.449 [-3.630, 0.732] | 2.213 | -.092 | -1.747 | .082 |
| MVPA (accelerometer-measured) x PA-related affect regulation (PA-AR) | 3.137 [-1.484, 7.759] | 4.689 | .085 | 1.476 | .141 |
| LPA (accelerometer-measured) x PA-related affect regulation (PA-AR) | -0.003 [-0.023, 0.016] | 0.020 | -.010 | -0.166 | .869 |
| PA self-reported (BSA-F) x PA-related affect regulation (PA-AR) | 0.709 [-2.395, 3.814] | 3.151 | .031 | 0.576 | .565 |
| *Psychological factors* | | | | | |
| repetitive negative thinking (PTQ) | 0.184 [0.047, 0.321] | 0.139 | .174 | 2.609 | **.010** |
| sleep quality (PSQI) | 0.683 [0.394, 0.973] | 0.294 | .247 | 4.693 | **<.001** |
| perceived stress (PSS) | 0.562 [0.186, 0.938] | 0.382 | .173 | 3.001 | **.003** |
| fear of corona virus (PAS) | 0.164 [-0.026, 0.354] | 0.193 | .093 | 1.808 | .072 |
| emotional regulation (DERS) | 0.101 [0.039, 0.163] | 0.063 | .229 | 3.385 | **.001** |
| emotional intelligence (TEIQue-SF) | -0.357 [-0.999, 0.286] | 0.652 | -.052 | -1.009 | .314 |

*Note.* Adjusted $R^2$=0.336; B: pooled unstandardized regression coefficient, 95% CI: 95% confident interval for B, SE: standard error, β: pooled standardized regression coefficient. Statistical significance: $p$<.05.
Exercise behavior: physical activity (PA; in minutes/week) assessed through accelerometer data and categorized by the metabolic equivalent of tasks (MET) into sedentary behavior (SB; <2 MET), light PA (LPA; 2.0-2.9 MET) and moderate to vigorous PA (MVPA; ≥3.0 MET) (Garber et al., 2011) or self-report via Physical Activity, Exercise, and Sport Questionnaire (BSA-F) (Fuchs, Klaperski, Gerber, & Seelig, 2015). PA-AR = PA-related affect regulation assessed with the corresponding subscale of the PA-related health competence questionnaire (Sudeck & Pfeifer, 2016).
Psychological factors: PTQ = Perseverative Thinking Questionnaire (Ehring et al., 2011); PSQI = Pittsburgh Sleep Quality Index (Buysse, Reynolds, Monk, Berman, & Kupfer, 1989); PSS = Perceived Stress Scale (Cohen, Kamarck, & Mermelstein, 1983; Klein et al., 2016); PAS = Pandemic Anxiety Scale (McElroy et al.,

2020); DERS = Difficulties in Emotion Regulation Scale (Gratz & Roemer, 2004; Gutzweiler & In-Albon, 2018); TEIQue-SF = Trait Emotional Intelligence Questionnaire – Short Form (Freudenthaler, Neubauer, Gabler, Scherl, & Rindermann, 2008; Petrides, 2009).
